# The values of coagulation function in COVID-19 patients

**DOI:** 10.1101/2020.04.25.20077842

**Authors:** Xin Jin, Yongwei Duan, Tengfei Bao, Junjuan Gu, Yawen Chen, Yuanyuan Li, Shi Mao, Yongfeng Chen, Wen Xie

## Abstract

**Objective:** To investigate the blood coagulation function in COVID-19 patients, and the correlation between coagulopathy and disease severity.

**Methods:** We retrospectively collected 147 clinically diagnosed COVID-19 patients at Wuhan Leishenshan Hospital of Hubei, China. We analyzed the coagulation function in COVID-19 patients through the data including thrombin-antithrombin complex (TAT), α2-plasmininhibitor-plasmin Complex (PIC), thrombomodulin (TM), t-PA/PAI-1 Complex (t-PAIC), prothrombin time (PT), international normalized ratio (INR), activated partial thromboplastin time (APTT), fibrinogen (FIB), thrombin time (TT), D-Dimer (DD), and platelet (PLT).

**Result:** The levels of TAT, PIC, TM, t-PAIC, PT, INR, FIB, and DD in COVID-19 patients were higher than health controls (p<0.05), and also higher in the patients with thrombotic disease than without thrombotic disease (p<0.05). What’s more, the patients with thrombotic disease had a higher case-fatality (p<0.05). TAT, PIC, TM, t-PAIC, PT, INR, APTT, FIB, DD, and PLT were also found correlated with disease severity. Meanwhile, we found that there were significant difference in TAT, TM, t-PAIC, PT, INR, APTT, DD, and PLT in the death and survival group. Further using univariate and multivariate logistic regression analysis also found that t-PAIC and DD were independent risk factors for death in patients and are excellent predicting the mortality risk of COVID-19.

**Conclusion:** The coagulation systems in COVID-19 patients are inordinate, and dynamic monitoring of them, might be a key in the control of COVID-19 death.

## Introduction

People are going through a battle against novel coronavirus pneumonia (COVID-19) all over the world. By 3 April 2020, more than 1,010,000 people have been infected and more than 50,000 have died worldwide. The 2019 novel coronavirus (SARS-CoV-2) was confirmed as the pathogen of the COVID-19 and belongs to Beta coronavirus by the phylogenetic analysis, which is similar to severe acute respiratory syndrome coronavirus (SARS-CoV) and middle east respiratory syndrome coronavirus (MERS-CoV). Respiratory symptoms, fever, dry cough, and panting, even acute respiratory distress syndrome (ARDS) and acute cardiac injury manifested in the COVID-19 patients [1-4].

SARS-CoV-2 can cause a series of disorders of the coagulation systems including endothelial damage, coagulation activation, and intravascular fibrin deposition. For COVID-19 patients in severe, coagulation activation can be leading to the formation of thrombus and even disseminated intravascular coagulation (DIC) [5]. And this coagulation activation is more common in patients with pre-existing coagulation disorders. On the other side, although, most severe COVID-19 patients treated with extracorporeal membrane oxygenation (ECMO) to temporarily replaces cardiopulmonary function, the coagulation disorder is still one of the most common complications of ECMO, which is also a common cause of disruption of ECMO [6-7]. Therefore, it is significant to monitor the coagulation systems parameters in COVID-19 patients to determine the coagulation function.

Previous studies have suggested that cytokine storms are typical abnormalities caused by SARS-CoV-2 [8-9]. And the underlying reason of the connection between the coagulation function and the disease severity was not completely clear. Especially, the index related thrombus such as thrombin-antithrombin complex (TAT), α2-plasmininhibitor-plasmin Complex (PIC), thrombomodulin (TM), t-PA/PAI-1 Complex (t-PAIC) are important markers in the process of venous thrombosis and significantly increased before thrombosis, which had not been reported. Therefore, this study was executed to evaluate the values of coagulation function for the prediction of severe COVID-19 patients.

## 2. Methods

### 2.1 Patients

We recruited 147 COVID patients and 18 health controls in Wuhan Leishenshan Hospital, Hubei, China. The patients were diagnosed and classified followed the New Coronavirus Pneumonia Prevention and Control Program (7th edition) published by the National Health Commission of China. Briefly, the controls were hospital employees who underwent the healthy examination. The study was approved by the Ethics Committee of Zhongnan Hospital of Wuhan University.

#### 2.1.1. Standard of severe patients

Severe patients should have any of the following conditions:

a. Respiratory distress, RR ≥ 30 times/minute;
b. Under the resting state, the oxygen saturation ≤ 93%;
c. Oxygen partial pressure (PaO2)/oxygen concentration (FiO2) in arterial blood ≤ 300 mmHg;
d. Lung imaging showed obvious progression of the lesion >50% within 24-48 hours.

#### 2.1.2. Standard of critical patients

Critical patients should have any of the following conditions:

a. Have respiratory failure and mechanical ventilation required;
b. Shock;
c. Complications of other organ failure require treatment in the ICU.

### 2.2 Coagulation system examination

Fasting whole blood from every subjects was collected in an EDTA or Sodium Citrate anticoagulant treated tube and analyzed within 30 minutes of collection. Coagulation systems parameters, thrombin-antithrombin complex (TAT), α2-plasmininhibitor-plasmin Complex (PIC), thrombomodulin (TM), t-PA/PAI-1 Complex (t-PAIC), prothrombin time (PT), international normalized ratio (INR), activated partial thromboplastin time (APTT), fibrinogen (FIB), thrombin time (TT), D-Dimer (DD), and platelet (PLT) were analyzed.

### 2.3 Statistical analysis

Statistical analysis was performed with SPSS version 25.0 software and Graphpad Prism (version 7.0). All the measurement data were tested for normality, and non-normally distributed data were expressed as median (interquartile range), nonparametric Mann-Whitney test was used for comparison between the two groups. Univariate and multivariate analysis using logistic regression analysis: calculate the odds ratio and 95% confidence interval. The prediction of various indicators for prognosis COVID-19 patients was analyzed by the receiver operating characteristic (ROC) curves, and the area under the ROC curve (AUC) was measured to evaluate the predictive ability. For all statistical analysis, P<0.05 was considered statistically significant.

## 3. Result

### 3.1 Clinical Characteristics

Among the 147 COVID-19 patients, 76 were males and 71 were females, the median age was 64 years, while in the control group, 14 subjects were male and 4 subjects were females (Table 1). The age was not significantly different among groups. There were 105 mild patients, 18 severe patients and 24 critical patients, of which 37 patients with thrombotic disease and 110 patients without thrombotic disease.

**Table 1.** Comparison of coagulation parameters between COVID-19 patients and Healthy control

|  | Healthy control (n=18) | COVID-19 patients (n=147) | P value |
| --- | --- | --- | --- |
| Age, median (IQR), y | 43 (34-51) | 64 (54-73) |  |
| Gender (Female/Male) | 4/14 | 71/76 |  |
| TAT | 1.70 (0.90-2.50) | 5.4 (2.9-14.5) | <0.001 |
| PIC | 0.41 (0.27-0.51) | 0.70 (0.54-1.10) | <0.001 |
| TM | 7.85 (6.90-9.18) | 10.00 (8.20-15.20) | <0.001 |
| t-PAIC | 5.95 (3.98-8.18) | 10.80 (7.10-14.85) | <0.001 |
| PT | 10.9 (10.48-11.33) | 11.30 (10.70-12.50) | 0.021 |
| INR | 0.93 (0.90-0.97) | 0.97 (0.92-1.08) | 0.019 |
| APTT | 27.15 (25.23-29.13) | 28.50 (24.95-32.80) | 0.105 |
| FIB | 2.71 (2.48-2.98) | 3.00 (2.62-3.90) | 0.029 |
| TT | 17.55 (17.08-18.13) | 17.30 (16.65-18.05) | 0.252 |
| DD | 0.15 (0.12-0.24) | 0.72 (0.29-2.29) | <0.001 |
| PLT | 270.50 (201.25-290.00) | 214.00 (178.50-265.5) | 0.083 |

Abbreviations: IQR, interquartile range. TAT, Thrombin-Antithrombin complex; PIC, α2-plasmininhibitor-plasmin Complex; TM, Thrombomodulin; t-PAIC, t-PA/PAI-1 Complex; PT, prothrombin time; INR, international normalized ratio; APTT, activated partial thromboplastin time; FIB, fibrinogen; TT, thrombin time; DD, D-Dimer; PLT, platelet.

### 3.2 Coagulation parameters of health controls and COVID-19 patients

The levels of coagulation parameters at health controls and COVID-19 patients were demonstrated in Table 1. COVID-19 patients had significantly higher values of TAT, PIC, TM, t-PAIC, PT, INR, FIB, and DD than health controls, and there were no significant differences in APTT, TT, and PLT.

### 3.3 Coagulation parameters of COVID-19 patients with thrombotic diseases vs patients without thrombotic diseases

Among the 147 patients, 37 patients with thrombotic disease, 110 were without the thrombotic disease. There were 7 patients died among the thrombotic disease group, while 3 patients died in the non-thrombotic disease group (Table 2). The patients in thrombotic disease group had a higher case-fatality rate than the non-thrombotic disease group (p<0.05). The levels of coagulation parameters at the thrombotic disease group and non-thrombotic disease group were demonstrated in Fig 1. COVID-19 patients with thrombotic disease had significantly higher levels of TAT, PIC, TM, t-PAIC, PT, INR, FIB, and DD than those without the thrombotic disease. There were no significant differences in levels of APTT, TT, and PLT.

**Table 2.** Case-fatality rate in the thrombotic disease and non-thrombotic disease

|  | Death | Survival | Total | Case-fatality rate |
| --- | --- | --- | --- | --- |
| Thrombotic disease | 7 | 30 | 37 | 18.9% |
| Non-thrombotic disease | 3 | 107 | 110 | 2.7% |
| Total | 10 | 137 | 147 | 6.8% |

**Fig 1.**
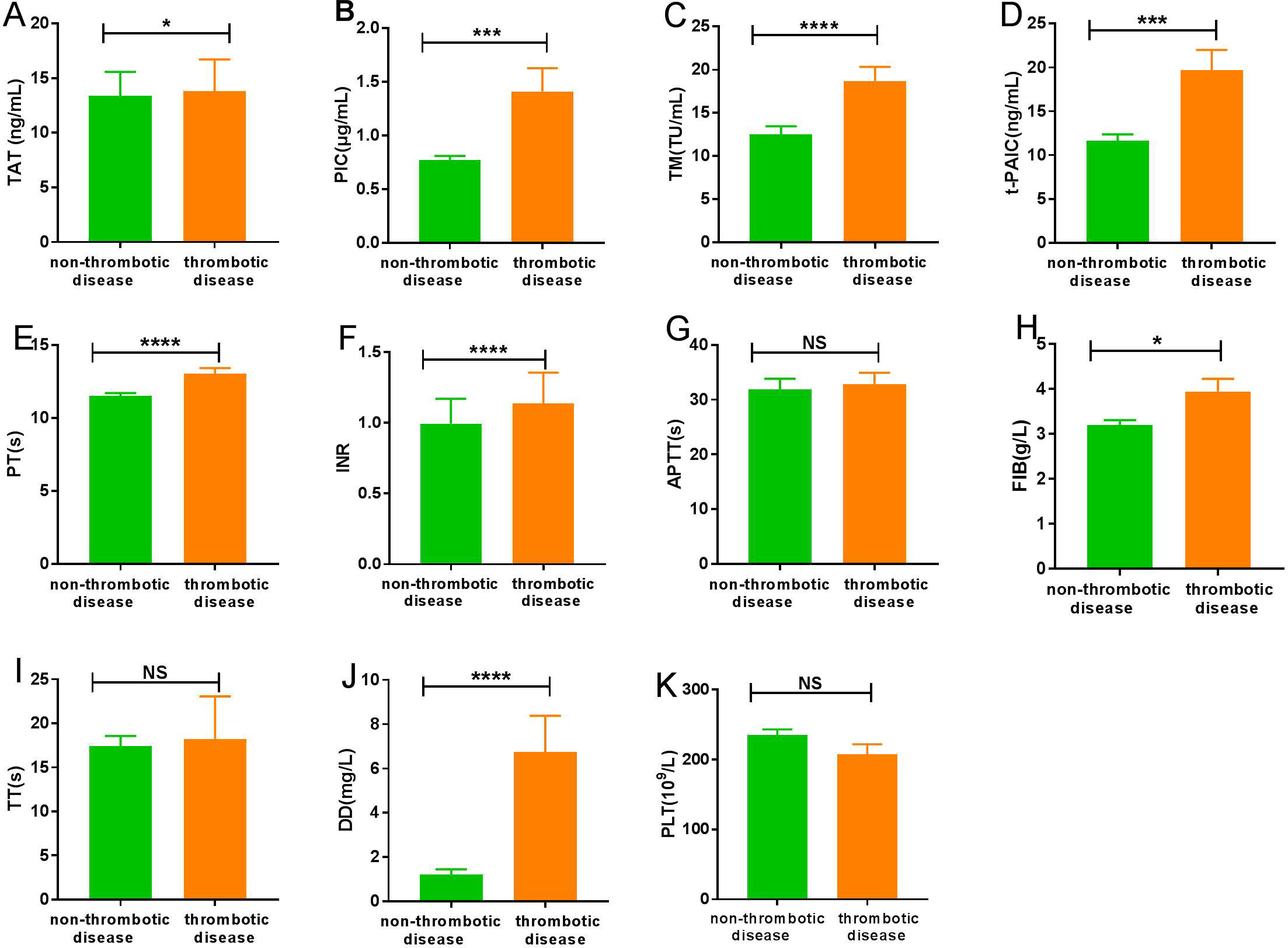
Characteristics of coagulation parameters for COVID-19 patients in non-thrombotic disease group and thrombotic disease group. (A) TAT; (B) PIC; (C) TM; (D) t-PAIC; (E) PT; (F) INR; (G) APTT; (H) FIB; (I) TT; (J) DD; (K) PLT. *P<0.05; ***P<0.001; ****P<0.0001; NS: P>0.05.

### 3.4 Coagulation parameters of three subgroups patients

Among the 147 included patients, 105 patients belonged to mild group, 18 were severe, and 24 were critical. TM, t-PAIC, PT, INR, DD were significantly different among the three groups, while TT was no significant difference (Figs 2C-F, I, J). The more severe in COVID-19 patients, the higher the levels of TM, t-PAIC, PT, INR, and DD. TAT and PLT were significantly different between the severe and critical groups or mild and critical group, and no significance were found between the mild and severe groups (Figs 2A, K). With regard to PIC and APTT, there were significant difference between mild and severe or mild and critical patients, and no significant difference in severe and critical group (Figs 2B, G). In addition, there was a significant difference that the level of FIB in severe group and mild group, however no significant difference was found between severe and critical group or mild and critical patients (Fig 2H). The ROC curves and AUC (Fig 3) demonstrated that TM (AUC=0.85), t-PAIC (AUC=0.84), PT (AUC=0.90), DD (AUC=0.92) had good predictive ability in mild VS severe/critical (all p<0.05, Fig 3).

**Fig 2.**
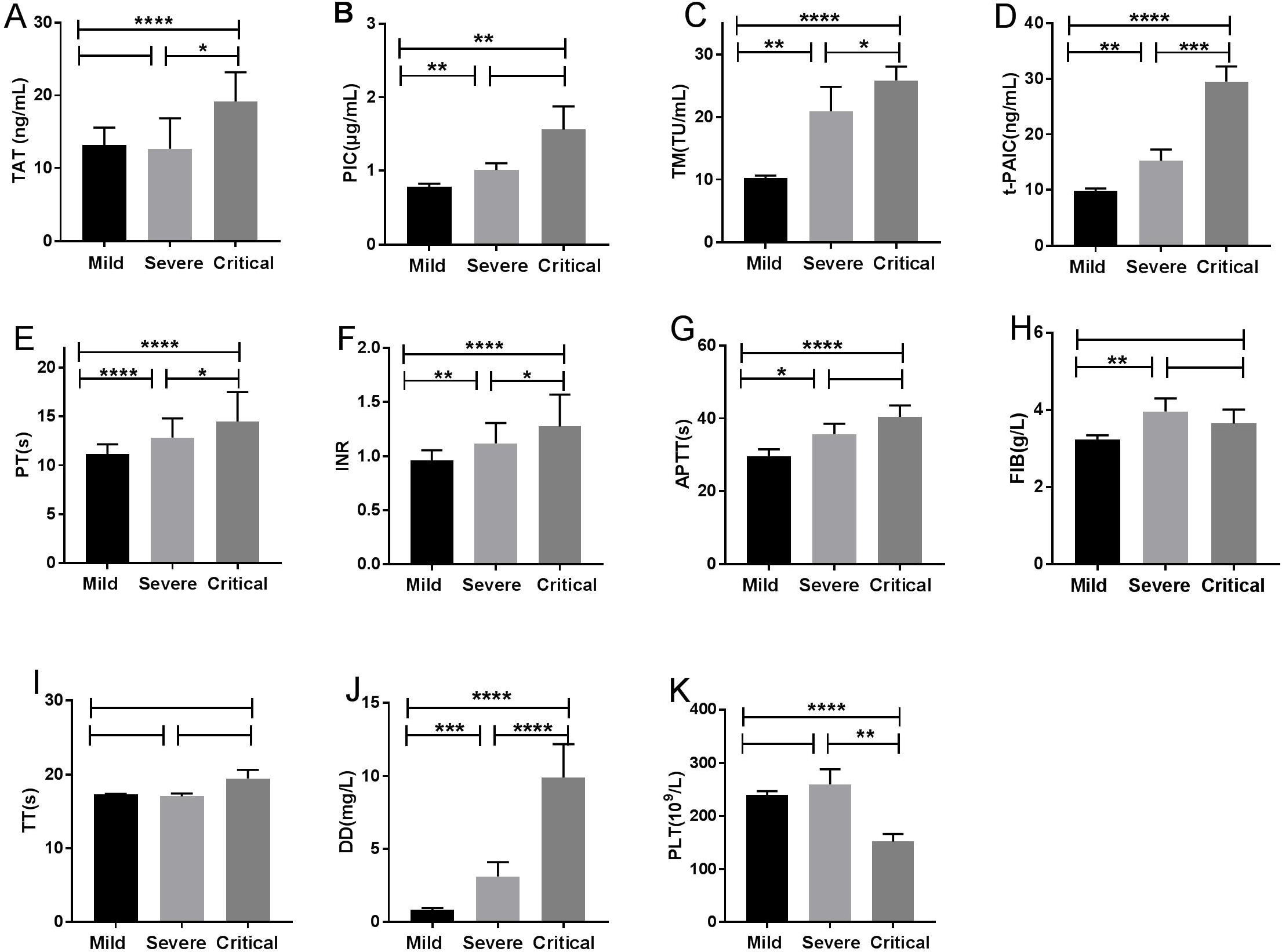
Characteristics of coagulation parameters among mild, severe and critical patients with COVID-19 pneumonia. (A) TAT; (B) PIC; (C) TM; (D) t-PAIC; (E) PT; (F) INR; (G) APTT; (H) FIB; (I) TT; (J) DD; (K) PLT. *P<0.05; **P<0.05; ***P<0.001; ****P<0.0001; NS: P>0.05.

**Fig 3.**
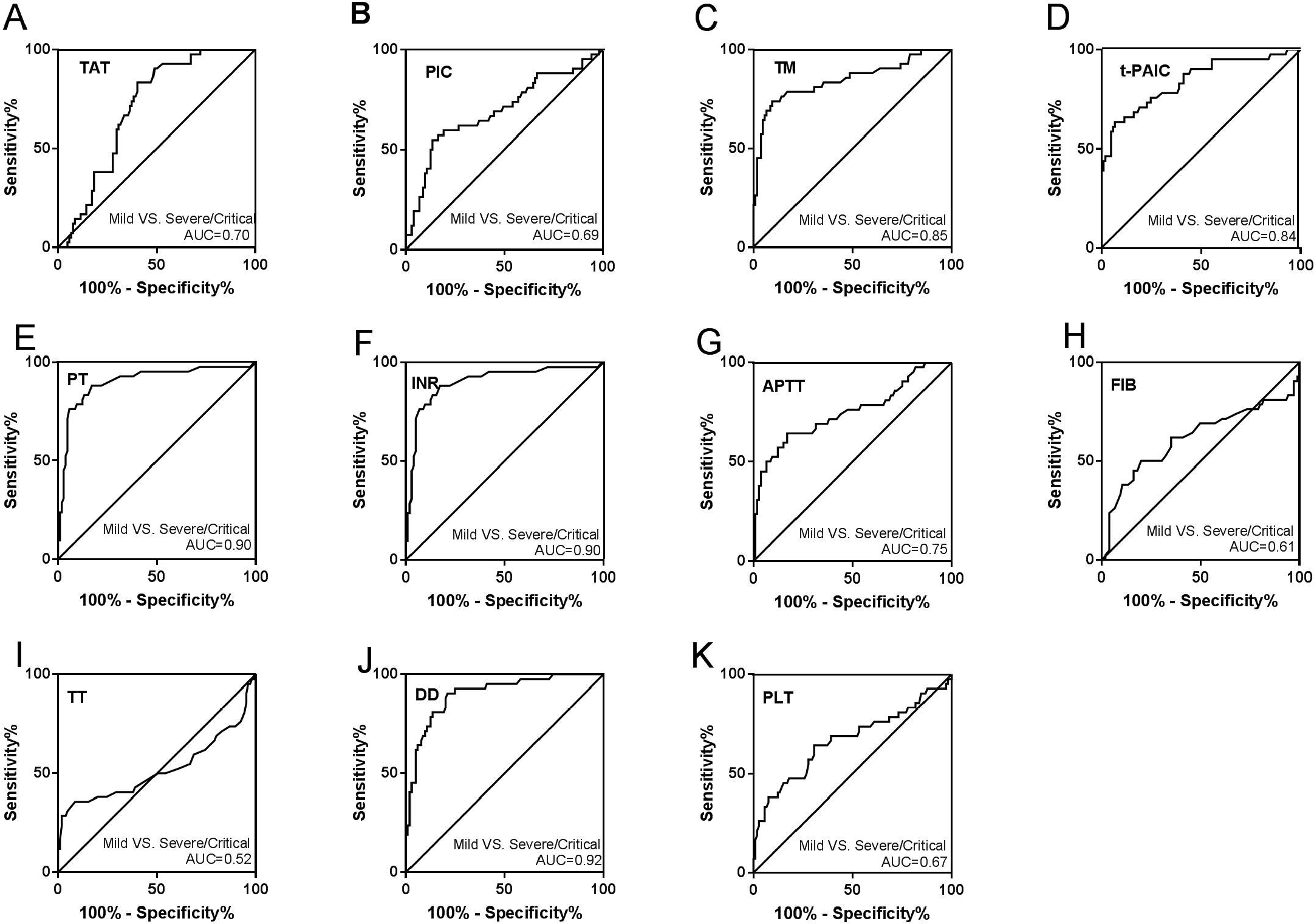
Receiver operator characteristic curves for coagulation parameters of COVID-19 patients in mild VS severe/critical. (A) TAT; (B) PIC; (C) TM; (D) t-PAIC; (E) PT; (F) INR; (G) APTT; (H) FIB; (I) TT; (J) DD; (K) PLT.

### 3.5 Coagulation parameters of COVID-19 patients in death and survival group

The levels of coagulation parameters at death and survival group were demonstrated in Fig 4. COVID-19 patients in the death group had significantly higher levels of TAT, TM, t-PAIC, PT, INR, APTT, and DD, but lower PLT levels than the survival group. There were no significant differences in levels of TT, PIC, and FIB. Further analysis using univariate and multivariate logistic regression also found that t-PAIC and DD were independent risk factors for patients death (Table 3). ROC were drawn to analyze the prognostic values of coagulation parameters in COVID-19 patients, the AUC of t-PAIC and DD were 0.92 [95% confidence interval (95%CI)=0.92-1.01], 0.94 [95%CI=0.90-0.98] (Fig 5). It was showed that the t-PAIC and DD are excellent in independently predicting the mortality risk of COVID-19. The sensitivity and specificity of t-PAIC in predicting death respectively were 90.0% and 91.2% with the cut-off of greater than 20.75 ng/mL; and those for DD were 100.0% and 86.0% with the cut-off of greater than 2.79 mg/L.

**Table 3.**
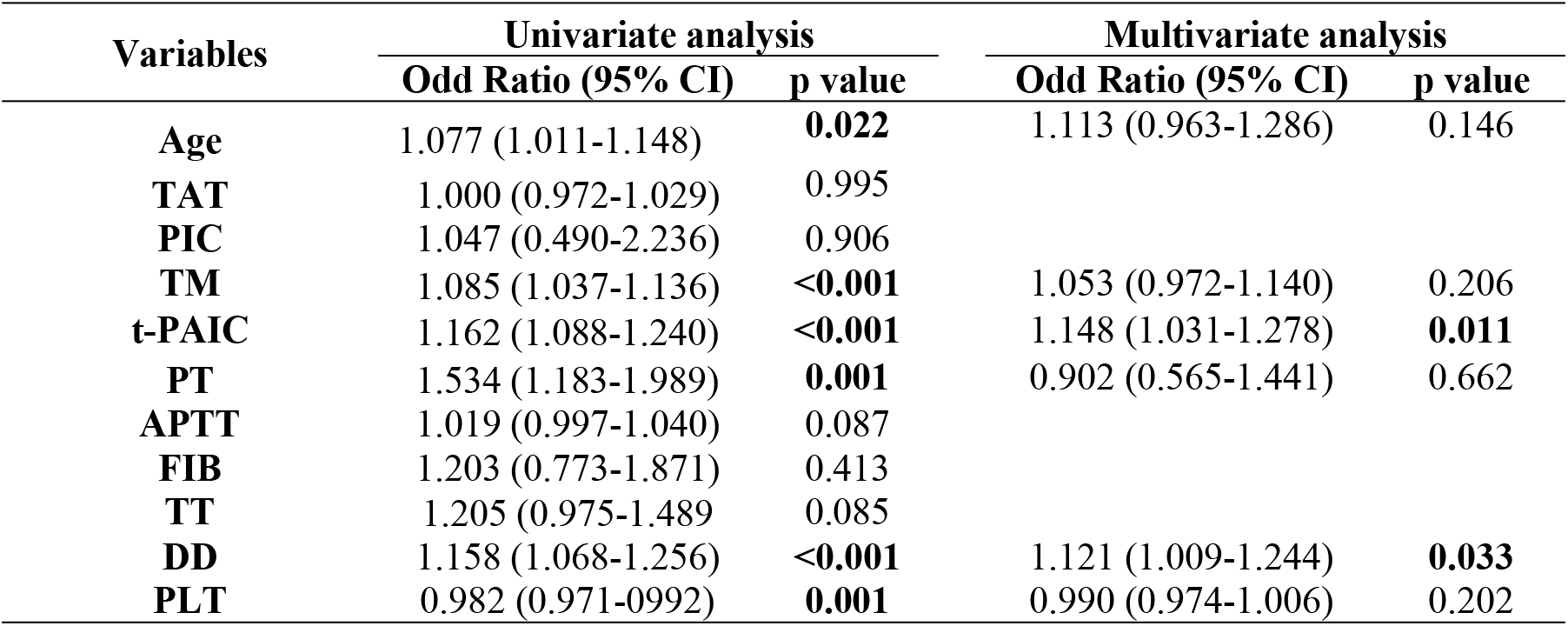
Univariate and Multivariate logistic regression analyses for factors predictive of COVID-19 death.

| Variables | Univariate analysis |  | Multivariate analysis |  |
| --- | --- | --- | --- | --- |
|  | Odd Ratio (95% CI) | p value | Odd Ratio (95% CI) | p value |
| Age | 1.077 (1.011-1.148) | 0.022 | 1.113 (0.963-1.286) | 0.146 |
| TAT | 1.000 (0.972-1.029) | 0.995 |  |  |
| PIC | 1.047 (0.490-2.236) | 0.906 |  |  |
| TM | 1.085 (1.037-1.136) | <0.001 | 1.053 (0.972-1.140) | 0.206 |
| t-PAIC | 1.162 (1.088-1.240) | <0.001 | 1.148 (1.031-1.278) | 0.011 |
| PT | 1.534 (1.183-1.989) | 0.001 | 0.902 (0.565-1.441) | 0.662 |
| APTT | 1.019 (0.997-1.040) | 0.087 |  |  |
| FIB | 1.203 (0.773-1.871) | 0.413 |  |  |
| TT | 1.205 (0.975-1.489) | 0.085 |  |  |
| DD | 1.158 (1.068-1.256) | <0.001 | 1.121 (1.009-1.244) | 0.033 |
| PLT | 0.982 (0.971-0.992) | 0.001 | 0.990 (0.974-1.006) | 0.202 |

**Fig 4.**
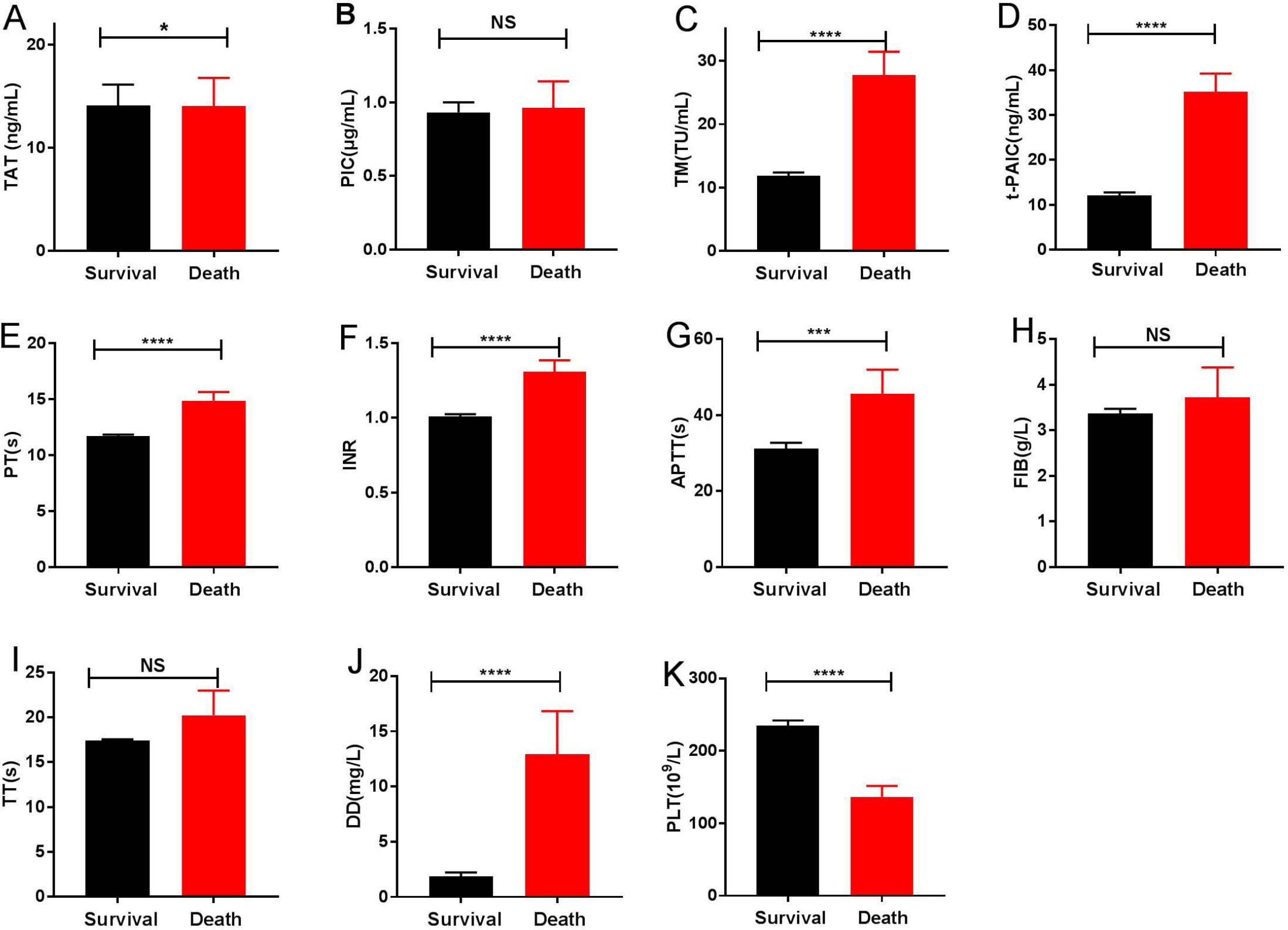
Characteristics of coagulation parameters in death and survival group. (A) TAT; (B) PIC; (C) TM; (D) t-PAIC; (E) PT; (F) INR; (G) APTT; (H) FIB; (I) TT; (J) DD; (K) PLT. *P<0.05; ***P<0.001; ****P<0.0001; NS: P>0.05.

**Fig 5.**
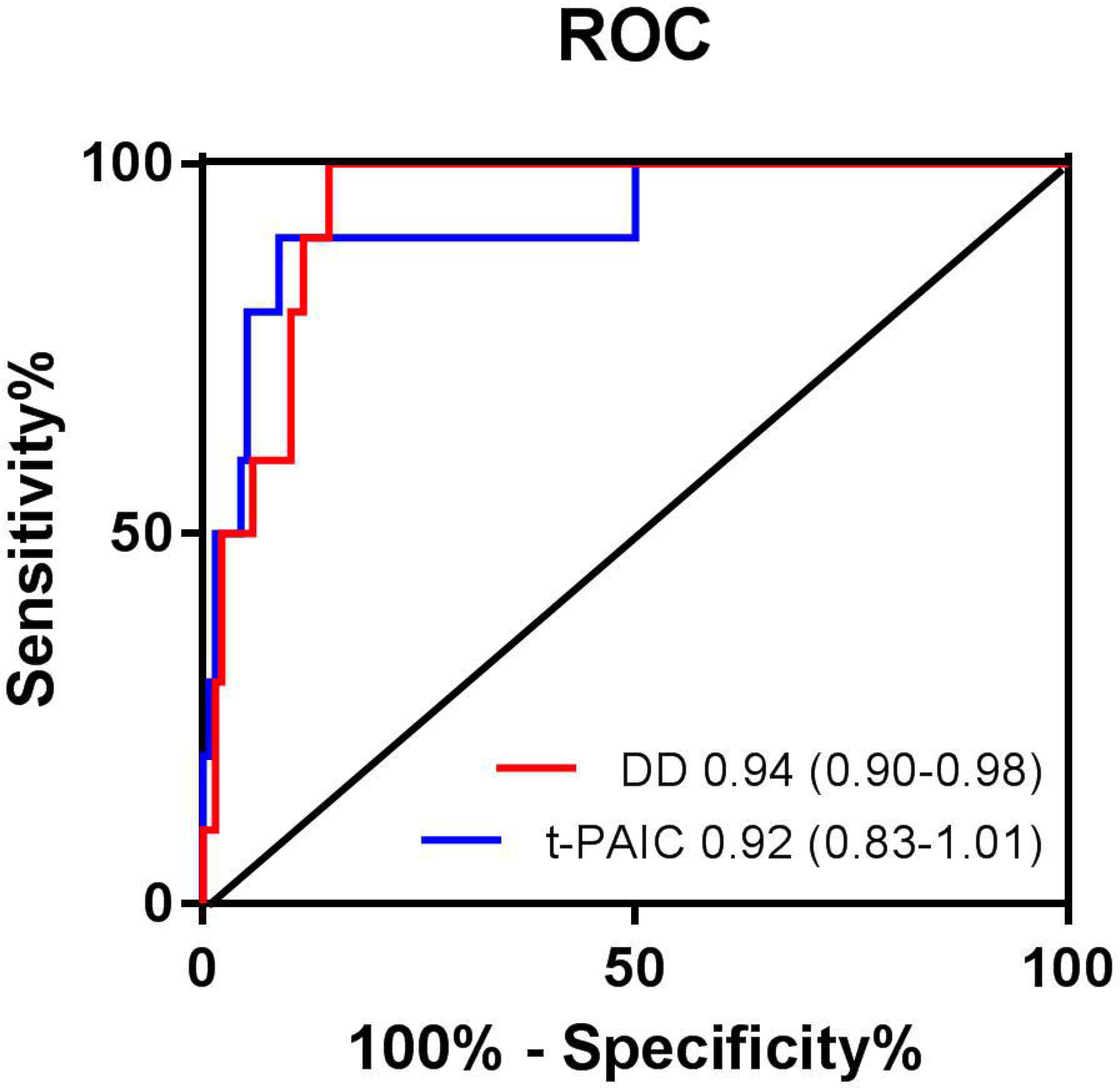
Receiver operator characteristics curves for coagulation parameters prediction of COVID-19 prognosis.

## 4. Discussion

We reported the coagulation function of COVID-19 patients in Wuhan Leishenshan Hospital of Hubei, China. The world health organization had declared the COVID-19 was an international public health emergency at the end of January [10]. Within a period of four months, more than 1,010,000 people have been infected and more than 50,000 people have died worldwide. Although most of the COVID-19 patients have mild symptoms with good prognosis and the crude mortality rate was about 2.3%, for patients progressing to severe or critical, mortality rates have increased significantly with crude death rates reaching 49% for critically [11-12]. Early identification of severe patients will help improve recovery rates and reduce mortality.

Studies have shown that COVID-19 patients can develop a variety of secondary diseases that cause pathological changes such as hematology, immunity, biochemistry, etc. [13-14], but the changes in coagulopathy are not well understood. In this study, a retrospective analysis was conducted about the coagulation systems parameters of 147 patients. TAT, PIC, TM, t-PAIC, PT, INR, APTT, FIB, TT, DD, and PLT are expected to assess the function of the coagulation systems in patients and indicate the severity of the patients to provide clinical assistance.

COVID-19, as a life-threatening infectious disease, can cause endothelial damage, coagulation activation, and intravascular fibrin deposition. Results of this study indicated that compared with the health controls, the TAT, PIC, TM, t-PAIC, PT, INR, FIB, and DD higher in COVID-19, while the APTT, TT, and PLT with no difference. Meanwhile, they were also no significant differences in thrombotic disease group and the non-thrombotic disease group. While the levels of TAT, PIC, TM, t-PAIC, PT, INR, FIB, and DD in thrombotic disease group were higher than non-thrombotic disease group. Four items of thrombosis detection (TAT, PIC, TM and t-PAIC) can predict the possibility of thrombosis in the patients at an early stage. TAT as a marker for activation of the coagulation systems, to determine the optimal period of anticoagulation therapy and early diagnosis of thrombosis and pre-DIC [15-16]. PIC as a marker of fibrinolytic system activation, predicting the formation of thrombus, assisting the diagnosis of DIC, and guiding antifibrinolytic treatment [17]. The elevated level of TM indicates an impaired vascular endothelial system [18-19]. t-PAIC, key marker of fibrinolytic system, suggesting thrombus progression [20-21]. Plasma levels of these sensitive biomarkers of endothelial injury in COVID-19 patients may be useful for evaluating the endothelial injury in COVID-19. Besides, we also found the COVID-19 patients with thrombotic disease had a higher case-fatality rate. Therefore, it is a situation that requires our vigilance when the coagulation systems parameters are abnormal in COVID-19 patient, and it is likely to be accompanied by thrombosis, and the risk of death is greater.

TAT, PIC, TM, t-PAIC, PT, INR, APTT, FIB, DD, and PLT were also found correlated with disease severity, while the TT was no correlation (Fig 2). TT is the time for plasma to coagulate after adding a prothrombin solution in the plasma, and reflects the presence of anticoagulants. Based on clinical practice and ROC analysis between mild and non-mild patients (Fig 3), some cut-off values of the test items were obtained. With TM >13.65 TU/mL, t-PAIC >15.25 ng/mL, PT >11.55 s, DD>1.03 mg/L, progress to severe illness should be closely observed and prevented. Among them, the rise of DD concentration is most obvious, similar to Wang’s [22] and Guan [23] study. DD is elevated because inflammation causes coagulation activation. Our results suggest the value of coagulation systems parameters in patients, and more attention about these to avoid the COVID-19 progression. As reported, anticoagulant treatment is required in severe and critical patients, and DD ≥ 5 µg/mL is used as an indicator to adjust anticoagulant therapy, while improving the prognosis [24]. To reverse intravascular coagulation, microthrombi formation, fibrin deposition, COVID-19 patients might need anticoagulant or fibrinolytic therapy.

Further, we found the COVID-19 patients in death group had significantly higher levels of TAT, TM, t-PAIC, PT, INR, APTT, and DD than the survival group, and the PLT decreased. It is shown that the patients have thrombocytopenia, elevated level of DD, and a prolonged APTT, suggesting that COVID-19 patients death may be associated with DIC [25-26]. Most severe COVID-19 patients develop dyspnea or hypoxemia one week after the onset, and their lung is more serious and gas exchange cannot be performed. When ventilator cannot restore lung function, ECMO has become an effective treatment to rescue critical patients, but it is only temporarily replacing cardiopulmonary function, and there are still many complications. Among them, the coagulation systems disorder is still one of the most common complications of ECMO, which terminate the ECMO [27-28]. Therefore, safe and scientific monitoring of the ECMO loop and thrombosis in patients, then early intervention can further reduce the harm of these complications to promote effective circulation support for ECMO. TAT, PIC, TM, and t-PAIC are the indicators of early prediction and monitoring of DIC, and DD is the indicator that a thrombus is occurring or ongoing. What’s more, our findings demonstrated that the prediction of the t-PAIC and DD can be as the excellent independently predicting the mortality risk of COVID-19, with an AUC of 0.92, 0.94, respectively (Fig 5).

Coagulation systems have values in COVID-19 patients because most patients have coagulopathy. Preventing recognition or blocking the occurrence of thrombus or DIC in COVID-19 patients, dynamic monitoring of coagulation systems parameters might be a key in the control of COVID-19 death.

## Corresponding Author

Correspondence and requests for materials should be addressed to Wen Xie.

^¶^ These authors contributed equally.

## Data Availability

We state all data referred to in the manuscript are available

## Author Contributions

Wen Xie had the idea for and designed the study. Wen Xie, Xin Jin and Yongwei Duan had full access to all of the data in the study and take responsibility for the integrity of the data and the accuracy of the data analysis.Wen Xie contributed to critical revision of the report. Xin Jin contributed to the statistical analysis. All authors contributed to data acquisition, data analysis, or data interpretation, and reviewed and approved the final version. Xin Jin and Yongwei Duan contributed equally and share first authorship. Xin Jin and Yongwei Duan contributed equally to this article.

### Acknowledgements

We acknowledge all health-care workers involved in the diagnosis and treatment of patients in Wuhan.

## Conflicts of Interest

The authors declare no conflict of interest.

## Funding

This research did not receive any specific grant from funding agencies in the public, commercial, or not-for-profit sectors.

## Notes

### Competing Interest Statement

The authors have declared no competing interest.

